# Uncertainty reduction in logistic regressions: a COVID-19 case-study using surrogate locations’ asymptotic values

**DOI:** 10.1101/2020.12.14.20248184

**Authors:** Bruno Hebling Vieira, Nathalia Hanna Hiar, George Cunha Cardoso

**Affiliations:** Department of Physics, FFCLRP, University of São Paulo, Ribeirão Preto, 14040-901, Brazil; Department of Biology, FFCLRP, University of São Paulo, Ribeirão Preto, 14040-901, Brazil; InBrain Lab, FFCLRP, University of São Paulo, Ribeirão Preto, Brazil

**Keywords:** Decision-making, COVID-19, logistic functions, Richards’, logistic function, Generalized Logistic Function

## Abstract

Logistic regressions are subject to high uncertainty when the data are not past the inflection point. For example, for logistic regressions estimated with data up to or before inflection point, uncertainties in the upper asymptotic value *K* can be of the same order of magnitude of the population under analysis. This paper presents a method for uncertainty reduction in logistic regression using data from a surrogate logistic process. We illustrate the procedure using the Richards’ growth function (Generalized Logistic Function) to make predictions for COVID-19 evolution in Brazilian cities at stages before and during their epidemic inflection points. We constrain the logistic function regression with *K* calculated from selected surrogate international cities where the epidemic is clearly past its inflection point. Information gained with this constraint stabilizes the logistic regression, reducing the uncertainty in the curves’ parameters, including the rate of growth at the inflection point. The uncertainty is reduced even when the actual surrogate *K* is used just as an anchor to simulate different epidemic scenarios. Results predicted for COVID-19 trajectories within Brazil agree with actual data. These results suggest that in the absence of big data, a simple logistic regression may provide low uncertainty if surrogate cities have been identified for estimates of *K*, even if the specifics of the evolution in the surrogate cities are different. The method may be used for other logistic models and for other logistic processes in other areas such as economics and biology, if surrogate processes can be identified.

## 1. Introduction

In real-world epidemic situations, compartmental models, which are based on average observable parameters, often fail to predict epidemic peak timing, case rate and total number of cases [1]. Published COVID-19 predictions in 2020 make clear that compartmental models are inadequate to deal with an emerging epidemic in an unknown complex network [2]. One of the causes of failure of compartmental models, such as the Susceptible-Exposed-Infected-Recovered (SEIR), is the assumption of a homogeneous basic reproduction number (*R*_*o*_) and homogeneous population behavior [3]. Agent networks can model the propagation of disease, but that would require thorough epidemiological knowledge of both the pathogen and the network of agents [4, 5, 6]. Logistic models are an intermediate alternative that has been shown to represent epidemic evolution reasonably well [7, 8, 9].

Logistic models describe monotonic growth processes that start growing exponentially or sub-exponentially, reach an inflection point, and asymptotically saturate to a maximum value. Examples of logistic models with exponential growth include the Verhulst function and the Richards’ growth *function* [10, *8], also known as generalized logistic function* (GLF). Subexponential growth logistic models include the generalized Richards’ model [11, 12, 7], which is defined by its differential equation without an analytical solution. For example, the number of accumulated cases *C*(*t*) in an epidemic is a type of problem that has been described by logistic functions [7, 8, 11]. As time passes, the availability of susceptible agents decreases, and *C*(*t*) grows asymptotically to the limit *K*. This is the upper asymptotic value of the logistic model,often referred to as carrying capacity when addressing population growth. In epidemics, *K* is the maximum number of accumulated cases expected for the population under study. It depends on the pathogen transmission mechanism, the social network structure and the underlying contagion dynamics [1]. The key benefit of logistic models is their ability to make predictions with little insight of the underlying specifics of the process, as is often the case in real epidemic surges.

In epidemic problems, the characterization of the logistic model around its inflection point enables estimation of its time derivative. Its peak, on the other hand, informs the time of maximum epidemic growth and the corresponding maximum rate of new cases per day. Logistic models accommodate different choices for the logistic function, and Verhulst’s is the most traditional one. It has been used to predict the evolution of COVID-19 *C*(*t*) in countries, by finding the number of cases *C*(*t*_critical_) = *K/*2 at its inflection point, which is fed into a machine learning algorithm to predict further evolution of the epidemic curve [13]. The procedure works as long as the inflection point has been reached in the data. Likewise, the GLF, also known as Richards’ growth model, was used to successfully predict the evolution of COVID-19 of various localities, as long as the inflection point has been identified [7, 12].

However, logistic models overfit the data if regression is done before its inflection point has been reached [8, 14]. In the exponential growth stage for *C*(*t*), regressions for forecasts are an ill-posed problem that leads to unacceptable uncertainties in the predictions; even estimates of the final number of accumulated cases *K* at the end of the epidemic have uncertainties of the order of the total population under consideration [12]. This is unfortunate because one of the reasons for modeling is to estimate both the time and the number of cases at the peak of the epidemic, either for preparedness or for the study of different strategies to deal with the disease.

Here, we use independent estimates of the logistic model’s upper asymptote *K* to constrain the GLF, allowing for stable non-linear regression on data series that end before or around the inflection point. We use surrogate cities from the United States, Italy, the United Kingdom, and Spain where the inflection point of the first COVID-19 wave has been identified, to estimate plausible values for *K*, that we use to constrain the GLF to predict scenarios in Brazilian cities. We need at least one surrogate city as an anchoring point. The choice of the GLF is for convenience, since it has been shown in the literature to work well for COVID-19 prediction [7]. To increase reliability of the data used, we use the accumulated fatalities count curve *D*(*t*), and estimate *C*(*t*) using the infection fatality rate (IFR). Our predictions use least squares curve-fitting of the constrained GLF on the cumulative fatalities curve data *D*(*t*), for different plausible values of *K*, for each Brazilian city under study.

This paper uses COVID-19 data to demonstrate the proposed strategy of using the asymptotic value of surrogate cities as an anchoring point for the development of scenarios of logistic growth in cities of interest. Our key contribution in this paper is offering an accessible early-predictions tool to gauge ranges of plausibility of the epidemic peak timing and magnitude in late onset cities. Our methodology gives a relatively narrow range of optimistic and pessimistic scenarios, using only *C*(*t*) data available before the epidemic peak has happened in the city of interest. Applications of our proposed methodology includes estimates of growth curves of less connected cities and countries for any process that obey logistic growth, such as epidemics.

## 2. Methodology

### 2.1 Generalized logistic function

We have used the GLF to model the evolution of fatalities [8, 7]:

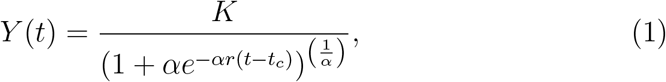

where *K* is the asymptotic value, *r* is the initial growth rate, *α* depends on the epidemic dynamics in the network, and *t*_*c*_ is the abscissa for the inflection point. The parameters *r* and *α* are known to trade-off such that *rα* equals the slope of the curve at the inflection point [15]. Two examples of Richards’ curves and their time derivatives are shown in Figure 1.

**Figure 1:**
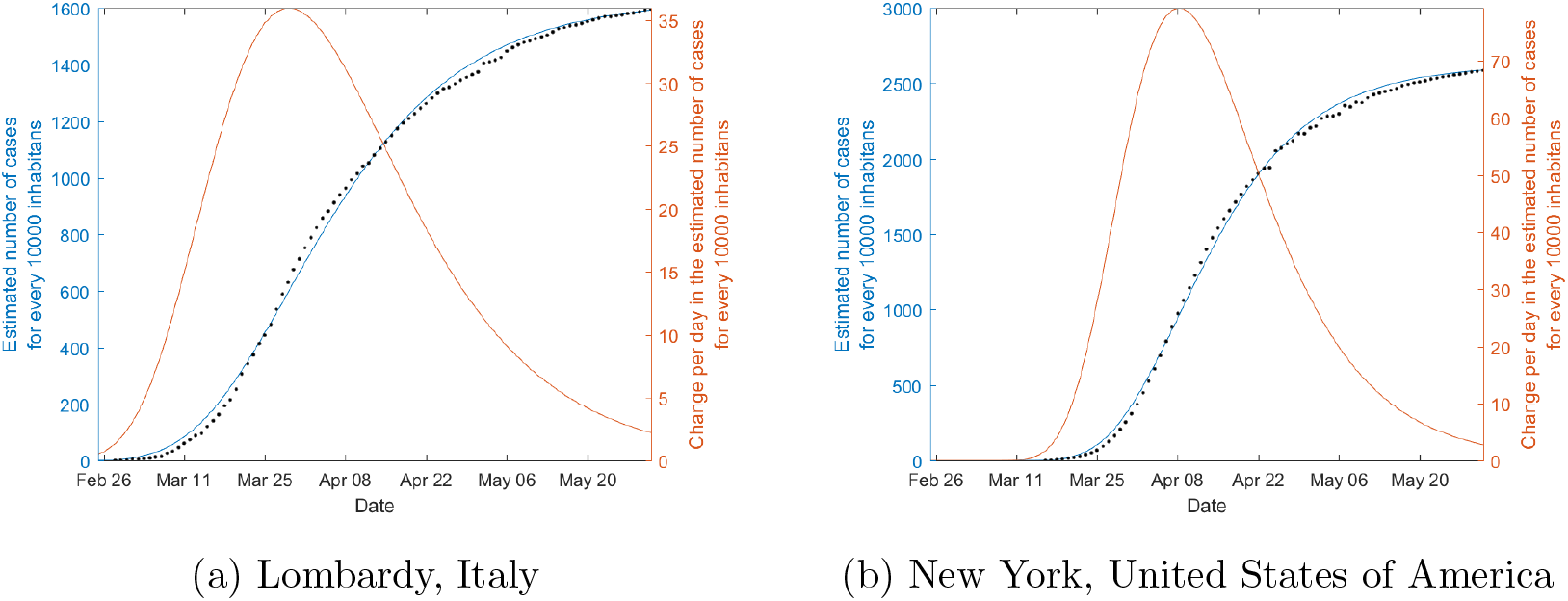
Examples of generalized logistic function regression to COVID-19 data of select regions, and respective time derivative. The proportion of fatalities divided by the IFR is shown in black dots.

To stabilize the logistic regressions of Equation 1 to data of cities of interest where the inflection point has not been reached yet, we will use *K* values derived from surrogate cities. The asymptote *K* for a city of interest could, in principle, be estimated from an appropriate epidemic model. For a system that obeys to the Susceptible-Infected-Recovered (SIR) model, the relationship between *R*_*o*_ and *K* is given by [16].

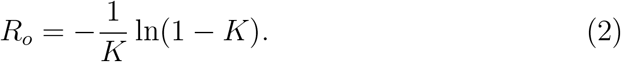

However, in most real-world scenarios compartmental models overestimates K especially for *R*_*o*_ with considerable heterogeneity [3] – which is a quite common occurrence. In addition, the relative uncertainty in *K* grows as *R*_*o*_ approaches one. More sophisticated models could be used for estimation of *K*, all suffering from inaccuracies due to lack of knowledge of the variance and higher moments of *R*_*o*_. Nevertheless, *R*_*o*_ may be used to establish an upper limit to *K*, irrespective of the existence of surrogate cities.

### 2.2. Data

We have analyzed fatalities data from select regions from the U.S.A., Italy, Spain, the U.K., and Belgium, which had gone through the peak in fatalities. We collected the data from the 2019 Novel Coronavirus Visual Dashboard operated by the Johns Hopkins University Center for Systems Science and Engineering (JHU CSSE; https://coronavirus.jhu.edu/), except for Queens borough, NY, USA (https://www.census.gov/quickfacts/fact/table/queenscountyqueensboroughnewyork/POP645218; https://www1.nyc.gov/site/doh/covid/covid-19-data-boroughs.page).

We have also selected fatalities data from cities in Brazil for analyses. (https://www.ibge.gov.br/cidades-e-estados; https://bigdata-api.fiocruz.br/relatorios). Details about the data and regions used can be found in https://github.com/bhvieira/SurrogateLocalitiesCovid.

For all regions, daily data comprised the period between the beginning of the epidemic and June/21/2020.

### 2.3. Modeling

A major issue with the early modeling of pandemic outbreaks is that the total capacity *K* in Equation 1 must be determined by data regression alone. This is a major source of uncertainty in the model. A possibility is using exponential growth modeling, thus forsaking the modeling of *K*, for early modeling [7]. We instead proposed to model different scenarios for a city based on plausible values for *K* derived by curve fitting *Y* (*t*) on surrogate cities data where the epidemic is clearly past the inflection point.

For the regression analyses we have used nonlinear least-squares optimization to estimate the free-parameters (*K, α, r, t*_*c*_) in Equation 1 for the surrogate cities, or (*α, r, t*_*c*_) for the cities of interest, where in the latter case *K* was kept fixed. Optimization was performed with a trust-region-reflective algorithm, implemented in the function “lsqcurvefit”, from the Optimization Toolbox in MATLAB. This optimizer, albeit more complex than traditional Levenberg-Marquardt based ones, can better cope with negative curvature of the estimate of Hessian of the objective (for a quick primer, see [17]). *α* and *r* are both constrained to the [0, 1] interval in the model, for stability [7]. Because *t*_*c*_ is not constrained, and will often be orders of magnitude larger than *α* and *r*, we divide its gradient by 10^3^. This is so because, otherwise, its gradient magnitude would dominate over other terms.

Since our objective function is non-convex on the parameters, one hundred random initialization were tried. Local minima initializations were not counted, but their resulting models remained on the pool of candidate models. Model selection was done by minimizing the Akaike information criterion. In our particular case, this resulted in picking the model with the highest likelihood, *i.e*. the one with the smallest mean squared error (MSE) given the Gaussian error assumption of nonlinear least-squares.

Instead of estimating our model from the actual cumulative number of reported cases *C*(*t*), we opted instead to estimate the number of cases from the cumulative number of fatalities *D*(*t*). Notoriously, the number of cases is susceptible to several testing and sampling biases. The number of fatalities *D*(*t*), while not completely free from its own biases, is more robust to them. We assume an IFR of 1% (See https://github.com/bhvieira/ SurrogateLocalitiesCovid for details on demographics age corrections made) and got the number of cases by dividing the number of fatalities by the IFR. This had the side effect of inducing a lag to our time estimates, since there is a time-shift between the cases and fatalities curve, such that *C*(*t − t*_*D*_) = *D*(*t*)*/*IRF, where *t*_*D*_ is the average time between diagnosis and death, in case of fatality. The delay *t*_*D*_ is also a source of uncertainty. To minimize this issue, we used *C*(*t − t*_*D*_) = *D*(*t*)*/*IRF for determination of *K* in surrogate cities and to illustrate *C*(*t*) in cities of interest, but use fatality rate *D*(*t*) directly in our estimations for *t*_*c*_ and the corresponding peak fatality rate.

### 2.4. Estimating the inflection point: the epidemic peak

In Equation 1, *t*_*c*_ is the time where the derivative 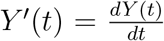 is maximal. From this, we estimate the actual peak in fatalities and its uncertainty. However, derivatives introduce uncertainty; to decrease uncertainty in *dY/dt*, we average the number of fatalities per day in an interval *T* approximately equal to the duration of the disease: *T* = 15 days around *t*_*c*_, as shown in Equation 3. The approximation to the nearest-integer is denoted by the *⌊ ·⌋* function.

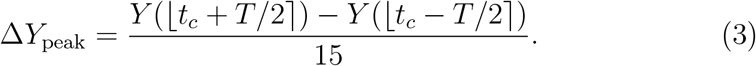

The uncertainty of Δ*Y*_peak_ can be approximated by the variance formula, resulting in Equation 4.

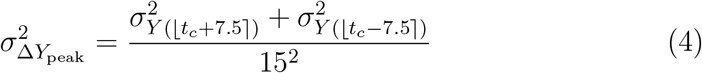

where the uncertainty 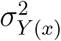 can be obtained by the formula for prediction intervals, using the conventional Delta method for uncertainty propagation.

## 3. Results

Results for *K* values for surrogate cities are shown in Table 1 for our data, corresponding to the first wave of COVID-19.

**Table 1:** Asymptotic logistic regression values for international localities. The pointestimated for *K* is shown alongside the 95% confidence interval between parentheses.

| Country | City | Asymptotic value (K) |
| --- | --- | --- |
| United States of America | Los Angeles | 0.0378 (0, 0.2180) |
|  | New York | 0.2617 (0.1326, 0.3907) |
|  | Rockland | 0.2011 (0, 0.4554) |
|  | Philadelphia | 0.1073 (0, 0.2853) |
|  | Fairfield | 0.1456 (0.0578, 0.2334) |
|  | Essex (MA) | 0.1434 (0.0377, 0.2491) |
| Italy | Lombardia | 0.1627 (0.0504, 0.2750) |
| Spain | Madrid | 0.2302 (0, 1) |
| United Kingdom | London | 0.0672 (0.0006, 0.1338) |
| Belgium | - | 0.0832 (0, 0.1683) |

Figure 2 shows GLF regressions for different fixed values of *K*, and also for *K* as a free parameter (automatic), for select Brazilian cities.

**Figure 2:**
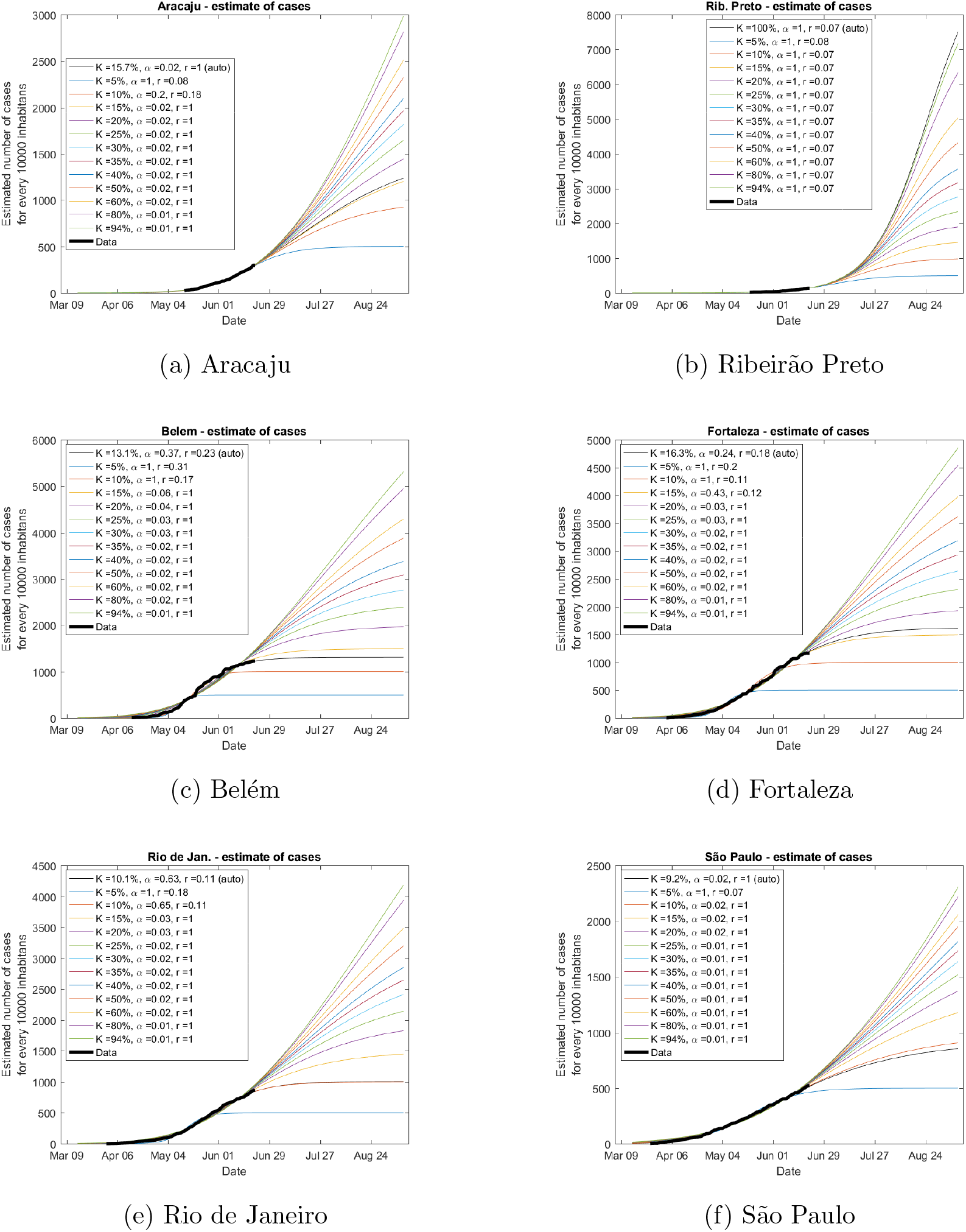
Prediction of epidemic evolution of the estimated number of cases for Brazilian cities. The thin black line denotes a model where the carrying capacity *K* was automatically optimized as a parameter (denoted “auto” in the legend).

Figure 3 shows the prediction of the day of the epidemic peak, given by *t*_*c*_ in Equation 1, and the prediction of the number of fatalities averaged in an interval *T* = 15 days centered on *t*_*c*_, as described in Equation 3.

**Figure 3:**
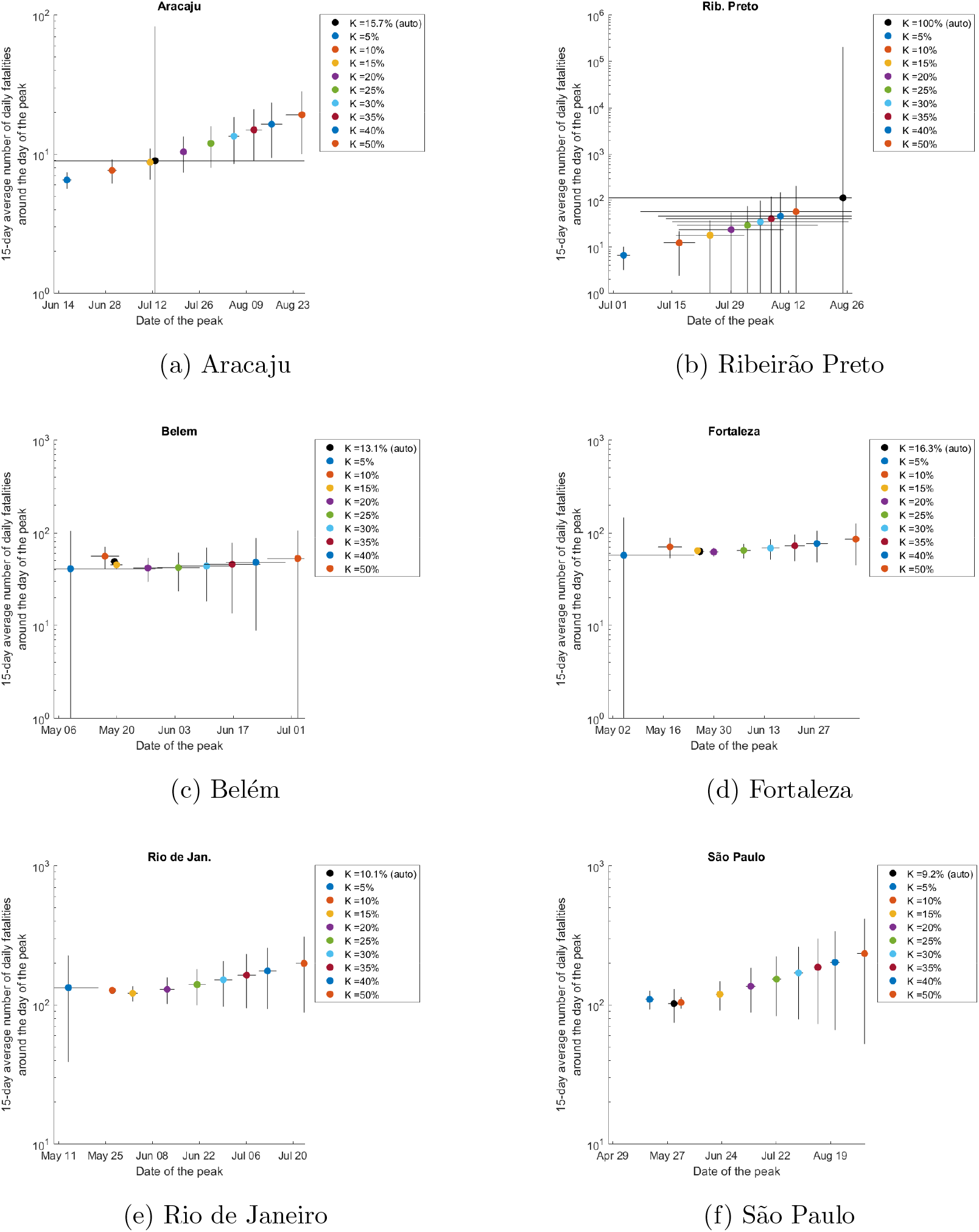
Prediction of the peak for Brazilian cities. The black dot (auto) represents a model where the carrying capacity *K* was automatically optimized as a parameter (denoted “auto” in the legend). To decrease uncertainty levels, results for the number of cases are averaged over 15 days centered on the peak day.

Figures for other cities studied are found in Table A.2 can be found in https://github.com/bhvieira/SurrogateLocalitiesCovid.

## 4. Discussion

The COVID-19 pandemic has revealed that sophisticated epidemic models, in the absence of granular agent-level knowledge of the disease, might not be enough to make good epidemic predictions [18]. Real-world epidemic outbreaks usually have poor quality data. Using the GLF has been shown to fit well the cumulative fatality curve in COVID-19 and other epidemics. However, when the total number of cases is not well past the inflection point, the GLF over-fits the data leading to uncertainties of the order of 100% [7]. Here we discuss the results of using the GLF to predict the evolution of cases *C*(*t*) in cities where the epidemic comes late. We use plausible ranges for the carrying capacity *K*, which are estimated from surrogate cities where the epidemic is beyond the inflection point. This alternative approach seeks to mitigate the over-fitting problem at the expense of increasing the uncertainty about the actual scenario that will develop in the city being forecast. Nevertheless, this strategy substantially decreases the range of predicted peak weeks and predicted peak number of cases per day, and decreases their statistical uncertainties compared to unconstrained fittings.

Using surrogate values for *K*, we performed GLF regression to Brazilian cities’ fatalities curves starting from the onset of COVID-19 until June, 21/2020. Comparison with the actual observed curve by October/2020, shows that the method gives ranges of expectation for the inflection point up to 100 times narrower than when *K* is left as a free parameter.

Here we will discuss the major results. In surrogate cities, where the pandemic had already passed the inflection point of the first wave, GLF regressions give *K* in the 5% to 25% interval (New York, London, Madri and other – see online material). For a Brazilian city still in the exponential phase of the epidemic (Figure 2a), we observe that the GLF regression with *K* as a free parameter leads to unacceptably high uncertainties (Figure 3a – auto). This high uncertainty is characteristic of a city where the inflection point of *C*(*t*) has not passed yet. The minimum possible value for *K* for Aracaju, given the data, is *∼* 10%, that represent the last data point in Figure 2a. Re-running the GLF regression with constrains 10% *< K <* 25%, the uncertainties are reduced (Figure 3a) and agree with actual observed data (July/2, 14 fatalities/day) [19], that is compatible with *K ∼* 12%. Notice the narrow predicted range, especially for the infection fatality rate, even if *K* = 25% is chosen.

Ribeirão Preto is another city shown still in the exponential phase (Figure 3b); when *K ∼* 12% is chosen, the predicted epidemic peak is compatible with the observed data (peak between July/7 and July/27, with *∼* 10fatalities at the peak [19]. Later we will discuss the choice for *K* = 12%, which is based on other Brazilian cities that are around or past the inflection point. Ribeirão Preto is a city where the epidemic curve is at the beginning. Notice that a tentative GLF regression leads to a carrying capacity of 100%, implying that the whole population would be infected at some point, which could not be correct because it would not take into account herd immunity. The uncertainties for Ribeirão Preto are higher than the ones for the other cities discussed in this paper because the data provided is at the very beginning of the exponential phase of the epidemic (Figure 2b).

Now let us look at Brazilian cities that had an earlier peak. These are probably more appropriate surrogates for other Brazilian cities, including the ones discussed above. Belém, Fortaleza, and Rio de Janeiro, Figure 2c, 2d and 2e, are past the epidemic peak in our time series, which is characterized in the GLF regressions by the small uncertainties in “auto”-fit for *K* (Figure 3c, *3d and 3e). The carrying capacity for the fist waves of Belém, Fortaleza and Rio de Janeiro are K ∼* 13%, *K ∼* 16%, and *K ∼* 10% respectively, which are compatible with the corresponding carrying capacities found for Aracaju and Ribeirão Preto, in the previous paragraph. We will now use 10% *< K <* 25% as a surrogate in the GLF regression to estimate the inflection point for the city of São Paulo.

The GLF regression for the city of São Paulo shows *K* = 9.2% when *K* is one of the free parameters (Figure 3f). Yet, the uncertainty in the inflection point’s position for *K* = 9.2% shows that the estimate may be improved. A carrying capacity of 9.3% would correspond to an epidemic peak at the end of May (in the past). For cities where the inflection point has happened in the past, the uncertainties were smaller. Let us use plausible values of K range from 10% to 16%, based on surrogate Brazilian cities. Figure 3f shows regressions with carrying capacities constrained to *K* = 10% and *K* = 15%, showing that the epidemic peak happened around the second week of June, which is compatible with official data for the complete fatality curve for São Paulo. The peak number of fatalities found is also compatible with 123 deaths/day for the 7-day moving average reported in the official data [19]. We can also observe in Figure 3f worse case scenarios, where *K* would grow to 25%. In such scenario the epidemic peak is predicted to happen a month later but with at least 50% more casualties — this could happen if the population that followed an initial trend relaxes their prophylactic measures.

It is important to have in mind the possibility of overloading the health system in a high *K* scenario. With overloading, it would be necessary to know the fraction of *C*(*t*) that need hospitalization, and what fraction of this number would convert into fatalities without hospitalization. Let us say, for example, that 12%*C*(*t*) needs hospitalization, and that without saturation of hospitals *D*(*t*) = 1%*C*(*t*). With saturation, a 10% increase in *C*(*t*) would raise fatalities to *D*(*t*) = (1% + 12%10%)*C*(*t*), which is more than a 100% increase in fatalities.

The method presented in this paper is useful in predictions for peripheral cities or countries with pandemics or epidemic-like phenomena, if an adequate surrogate city or country has been identified. As the pandemic evolves, cities geographically and socio-economically closer to the cities of interest can be used as surrogates, increasing the plausibility of adequacy of the surrogate *K*.

Our method may be compared to the one described in [15], where a Bayesian hierarchical model integrates multiple countries’ data for infection trajectory prediction of the spread of COVID-19 based on the Richards’ growth curve. The results reported, despite good rational and sophistication of the model [15], underestimated the number of cases in main COVID-19 countries by a factor of at least five, even if predictions were done after the inflection points had occurred for the countries of interest. Our method could suffer from a similar problem, especially if some Asian cities were used as surrogates for Brazilian cities. Many cities in Asia have government and population behavior that would not serve as surrogate for Brazilian cities’ management of COVID-19. It might become clear from the data that the asymptotic value *K* for Asian cities are not appropriate surrogates for a Brazilian city when *K*_*asian*_ *≈ C*(*t*)_*Brazilian*_ even when the Brazilian cities below their epidemic inflection point.

One challenge of our method is the identification of a proper surrogate city. The biggest source of uncertainty in the evaluation of the peak number of cases per day is the IFR. The IFR, in conditions where the health system is not yet saturated, we have used as surrogates the IFRs for Italian, Korean and American cities or regions, corrected for the demographics of Brazilian cities, and concluded that a 1% IRF is a plausible value (Supplementary Material).

Changes in adherence to prophylaxis, such as social distancing, mask usage and self-isolation due to case tracking & contact tracing, will alter the value of *K*. Our model is agnostic to such possible changes, modeling instead a static *K*. Thus, it is more adequate for short to intermediary timescales.

As presented, the method seem to be more appropriate for the first wave or for single wave epidemics in a city. The first wave is the most important because it will get the health system unprepared. Care must be taken because for an arbitrary epidemic case it is not known if the chosen logistic function will be adequate for the data regression. This uncertainty creates an unknown bias to confidence intervals reported. However, this issue is common to all mathematical models. An intrinsic limitation to the GLF used is regarding its inflection point: it is limited between *K/*2 (for *α* = 1) and *K/e* (for *α →* 0). Yet, the literature shows that GLF represents a good balance between number of free parameters and ability to describe well epidemic curves [8]. Indeterminacy of *α* vs. *r* remained unresolved in our study, but different from [14], here *K* is pre-determined, allowing for continuous transitions between curves for different *K*’s. Finally, we also observed that the product *αr* is proportional to the slope of the logistic function [15]. Because of the difficulty in determining both *α* and *r* independently, in some references *αr ∝*= *r*_*o*_, where *r*_*o*_ is not represent the initial growth of the process [8]. More sophisticated logistic models may be used for the constrained regression for the creation of alternative predictive scenarios, for example, using generalized exponentials [20, 21].

In conclusion, the proposed method gives a complementary prescription to the traditional growth prediction methods. It can apply to any early logistic process, where there is an earlier surrogate process for estimation of the carrying capacity. This strategy overcomes the intrinsic inability logistic models have for prediction of the inflection point. It can be used not only for epidemics but also for commerce, economics, viral information dissemination in a population. Our method lowers the uncertainty in the prediction for optimistic and pessimistic epidemic scenarios without the need for sophisticated models or big data type of resources, and gives decision-makers anchor points specific for their situation.

### Data and code availability statement

Code and data are available online. See: https://github.com/bhvieira/CovidRichards/

## Data Availability

All data referred to in the manuscript is made available in a public repository. See https://github.com/bhvieira/SurrogateLocalitiesCovid

https://www.ibge.gov.br/cidades-e-estados/

https://bigdata-api.fiocruz.br/relatorios/

https://www.census.gov/data/tables/time-series/demo/popest/2010s-total-cities-and-towns.html

https://github.com/CSSEGISandData/COVID-19/tree/master/csse_covid_19_data/csse_covid_19_time_series

http://demo.istat.it/bilmens2019gen/index.html

https://statistichecoronavirus.it/regioni-coronavirus-italia/lombardia/

http://www.madrid.org/iestadis/fijas/estructu/demograficas/padron/estructupopc.htm

https://www.comunidad.madrid/servicios/salud/2019-nuevo-coronavirus

https://www.citypopulation.de/en/uk/greaterlondon/

## Acknowledgments

We thank Professor Alexandre S. Martinez for critical reading of the manuscript.

## Appendix A Data from Brazilian and international cities

**Table A.2:** Data from representative Brazilian cities. Population and fatalities statistics for Brazil come from https://www.ibge.gov.br/cidades-e-estados/ and https://bigdata-api.fiocruz.br/relatorios/, respectively.

| City | Population | Date of first reported fatality |
| --- | --- | --- |
| Aracaju | 657013 | 02/04/2020 |
| Belém | 1492745 | 06/04/2020 |
| Brasília | 3015268 | 29/03/2020 |
| Campinas | 1204073 | 31/03/2020 |
| Curitiba | 1933105 | 06/04/2020 |
| Florianópolis | 500973 | 02/04/2020 |
| Fortaleza | 2669342 | 26/03/2020 |
| Joinville | 590466 | 30/03/2020 |
| Manaus | 2182763 | 30/03/2020 |
| Porto Alegre | 1483771 | 25/03/2020 |
| Recife | 1645727 | 25/03/2020 |
| Ribeirão Preto | 703293 | 26/03/2020 |
| Rio de Janeiro | 6718903 | 23/03/2020 |
| Salvador | 2872347 | 29/03/2020 |
| São Luís | 1101884 | 29/03/2020 |
| São Paulo | 12252023 | 17/03/2020 |
| Uberlândia | 691305 | 03/04/2020 |

**Table A.3:**
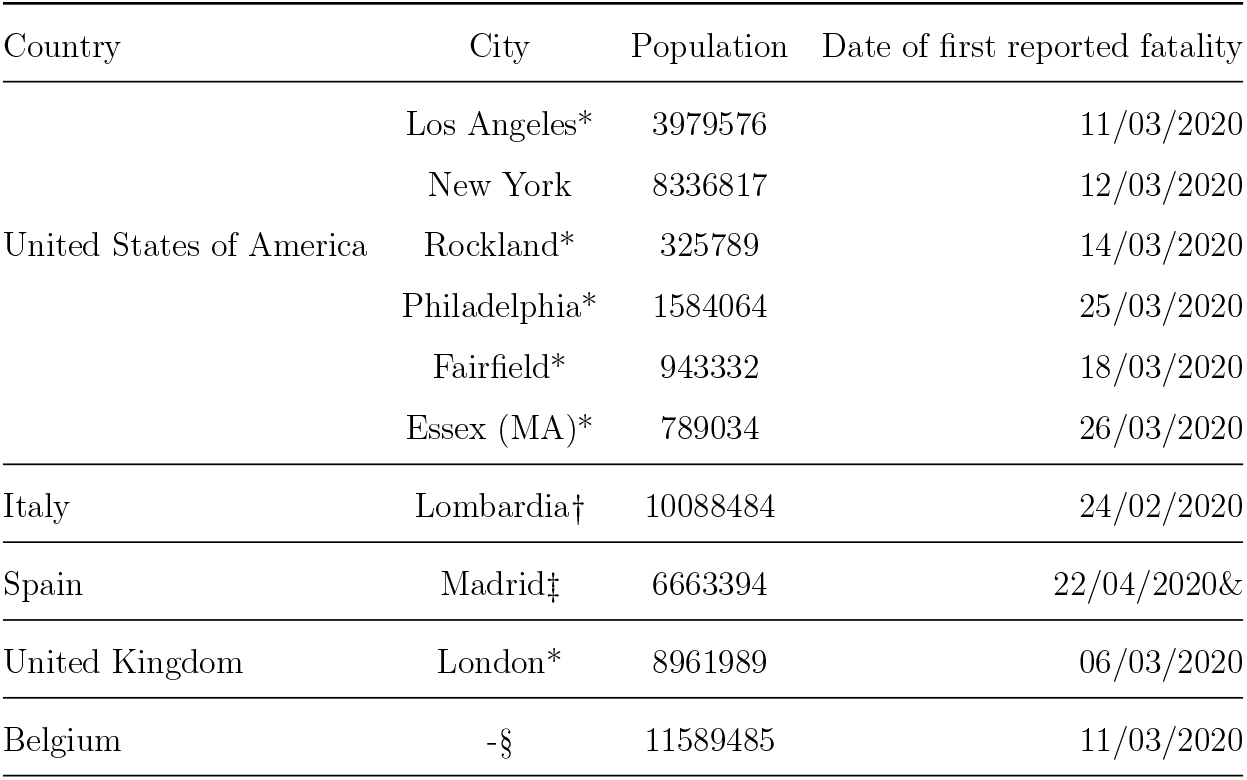
Data from representative international cities. *: county-level data; t: region-level data; t: Community-level data; §: country-level data; &: date report started. Population and fatalities statistics for the United States of America come from https://www.census.gov/data/tables/time-series/demo/popest/2010s-total-cities-and-towns.html and https://github.com/CSSEGISandData/COVID-19/tree/master/csse_covid_19_data/csse_covid_19_time_series, respectively. http://demo.istat.it/bilmens2019gen/index.html and https://statistichecoronavirus.it/regioni-coronavirus-italia/lombardia/forLombardia. http://www.madrid.org/iestadis/fijas/estructu/demograficas/padron/estructupopc.htm and https://www.comunidad.madrid/servicios/salud/2019-nuevo-coronavirus for Madrid. https://www.citypopulation.de/en/uk/greaterlondon/ and https://www.england.nhs.uk/statistics/statistical-work-areas/covid-19-daily-deaths/ for London. https://www.worldometers.info/world-population/belgium-population/ and https://coronavirus.jhu.edu/map.html for Belgium.

